# Health workers’ and administrators’ experiences of duty handover on maternity wards in public health facilities in Hoima District, Uganda: a qualitative phenomenological study

**DOI:** 10.64898/2026.10.21.26366155

**Authors:** Nicholas Tumwesige, Kawungu Saad Sessimba, Joshua Kitimbo, Herbert Kayiga, Mike N. Kagawa

## Abstract

**Background:** Duty handover is a high-risk point in the continuity of maternity care, and breakdowns in handover communication have been linked to delayed care and avoidable harm. Qualitative evidence from maternity wards in low-resource Sub-Saharan African settings is limited, and to our knowledge no study has examined handover experiences on maternity wards in Hoima District, Uganda. This study explored the experiences and challenges of health workers and administrators regarding duty handover on maternity wards in public health facilities in Hoima District, Uganda.

**Methods:** We conducted a qualitative phenomenological study. Fifteen in-depth interviews with maternity ward health workers and five key informant interviews with administrators were carried out across six public health facilities. Interviews were audio-recorded, transcribed verbatim and analysed using a modified Colaizzi seven-step method. Reflexivity and bracketing were addressed through a pre-fieldwork assumptions statement and bracketing interview, a reflexive journal kept throughout data collection, and independent double-coding of a subset of transcripts.

**Results:** Two themes were identified: duty handover practices on maternity wards, and challenges experienced by health workers during handover. Handover was inconsistent in structure, duration and adherence to protocol, and relied heavily on informal telephone communication rather than structured face-to-face reporting. Standardised tools were used in some facilities only. Participants identified time constraints and workload, inadequate training, limited resources and supervision, negative staff attitudes, and difficulty accessing patient information as key challenges, and linked communication breakdowns to delays and errors in care. Patients and families were largely not involved in handover.

**Conclusions:** Duty handover on the maternity wards studied was inconsistent and constrained by capability-, opportunity- and motivation-related barriers operating together. Standardised and locally adapted handover tools, targeted training, stronger supervision and facility- and national-level guidance appear necessary to protect continuity of care in comparable low-resource maternity settings.

## Introduction

Duty handover is the transfer of professional responsibility and accountability for some or all aspects of care for a patient, or group of patients, to another person or professional group on a temporary or permanent basis. It is one of the most vulnerable moments in the care continuum, and the World Health Organization has identified improved communication during patient handovers as a patient safety priority [1]. On maternity wards, where a woman’s or newborn’s condition can change within minutes, the information passed between shifts can affect whether deterioration is recognised and treated in time [2].

Maternal mortality remains concentrated in Sub-Saharan Africa, which accounted for roughly 70% of global maternal deaths in 2020 [3]. Uganda’s maternal mortality ratio is estimated at 189 deaths per 100,000 live births [4]. Communication gaps between shift teams have been shown to drive adverse care and treatment outcomes, yet analyses of the problem remain limited in low- and middle-income countries (LMICs) [5].

Handover has been studied extensively overall, including in systematic reviews of reviews [6], and structured communication tools such as SBAR and ISBAR (Introduction, Situation, Background, Assessment, Recommendation) are widely taught [7]. A recent prospective study in an Italian emergency department found that introducing a standardised ISBAR-based tool significantly improved the completeness and accuracy of information exchanged at shift change [8]. However, handover-improvement initiatives have mainly been tested in high-income countries, with very little work in LMICs [9].

The available African evidence describes handover that is largely informal and unstructured. In Kenyan newborn units, handover is mostly verbal at the bedside, formal reference documents are used rarely and ad hoc, nurses rely on personal notes and notebooks, and the content of handover varies within teams and across shifts [9,10]. In two referral hospitals in Ghana, communication gaps among shift teams were driven by poor documentation, limited training in handover communication and the absence of formal handover procedures [5]. In Irish maternity services, a qualitative descriptive study found a general absence of formal handover policy and training, with midwifery and medical handovers held separately [11], and a mixed-methods study of maternity shift changes in The Gambia examined handover communication in relation to women’s safety [2].

Patient and family involvement is a further dimension of patient-centred handover. Tailored ward-level interventions that bring handover to the bedside have been associated with greater patient participation and opportunities for patients to ask questions and raise concerns [12]. Participation in decision-making is, however, only partly institutionalised in maternal and newborn health systems across LMICs: an analysis of 102 national health ministries found that about half reported integrating community participation into national plans and only about one-third into implementation [13].

Anecdotal evidence from Maternal and Perinatal Death Surveillance and Response (MPDSR) reviews at the Hoima Regional Referral Hospital (HRRH) suggested that a substantial share of maternal and perinatal morbidity and mortality could be attributed, at least in part, to poor handover among health workers. We are not aware of a qualitative study that has explored handover on maternity wards in Uganda from the perspective of both frontline health workers and the administrators responsible for them.

This study, therefore, explored the experiences and challenges of health workers and administrators regarding duty handover on maternity wards in public health facilities in Hoima District, Uganda, applying the Capability, Opportunity, Motivation and Behaviour (COM-B) model to identify context-specific barriers and facilitators that could inform practical improvements.

## Materials and methods

### Ethics statement

The study was approved by the Makerere University School of Medicine Research Ethics Committee (Mak-SOMREC-2024-1081; approved at the 198th convened meeting on 10 December 2024, with approval valid from 10 March 2025 to 10 March 2026). Administrative clearance was obtained from the District Health Officer, the Hospital Director of HRRH and the in-charges of the participating health centres before data collection. All participants gave voluntary written informed consent before enrolment, after being told the purpose, procedures, risks and benefits of the study and the audio-recording procedures. They were informed that they could decline any question or withdraw at any time without consequence. No minors were enrolled. Confidentiality was protected using study codes and pseudonyms, and all data were stored securely.

### Study design

We used a qualitative phenomenological design [14]. Phenomenology was chosen because the study sought to understand how health workers and administrators experience, interpret and give meaning to duty handover, rather than to measure how often handover occurs or how closely it follows protocol. Data were analysed with Colaizzi’s descriptive phenomenological method [15], described below, so that the design and the analytic procedure are consistent.

### Conceptual framework

The study was guided by the Capability, Opportunity, Motivation and Behaviour (COM-B) model [16], which proposes that a behaviour—here, effective duty handover—is the product of interacting capability, opportunity and motivation (**Fig *1***). Capability covers knowledge of handover protocols, communication skills and competence in prioritising and transmitting clinical information; opportunity covers standardised tools, protected time, staffing and the physical environment; and motivation covers perceived importance of handover, accountability and attitudes to teamwork. The model informed interpretation of the findings at individual, interpersonal and systemic levels; themes were derived from the data through the phenomenological procedure described below.

**Fig 1.**
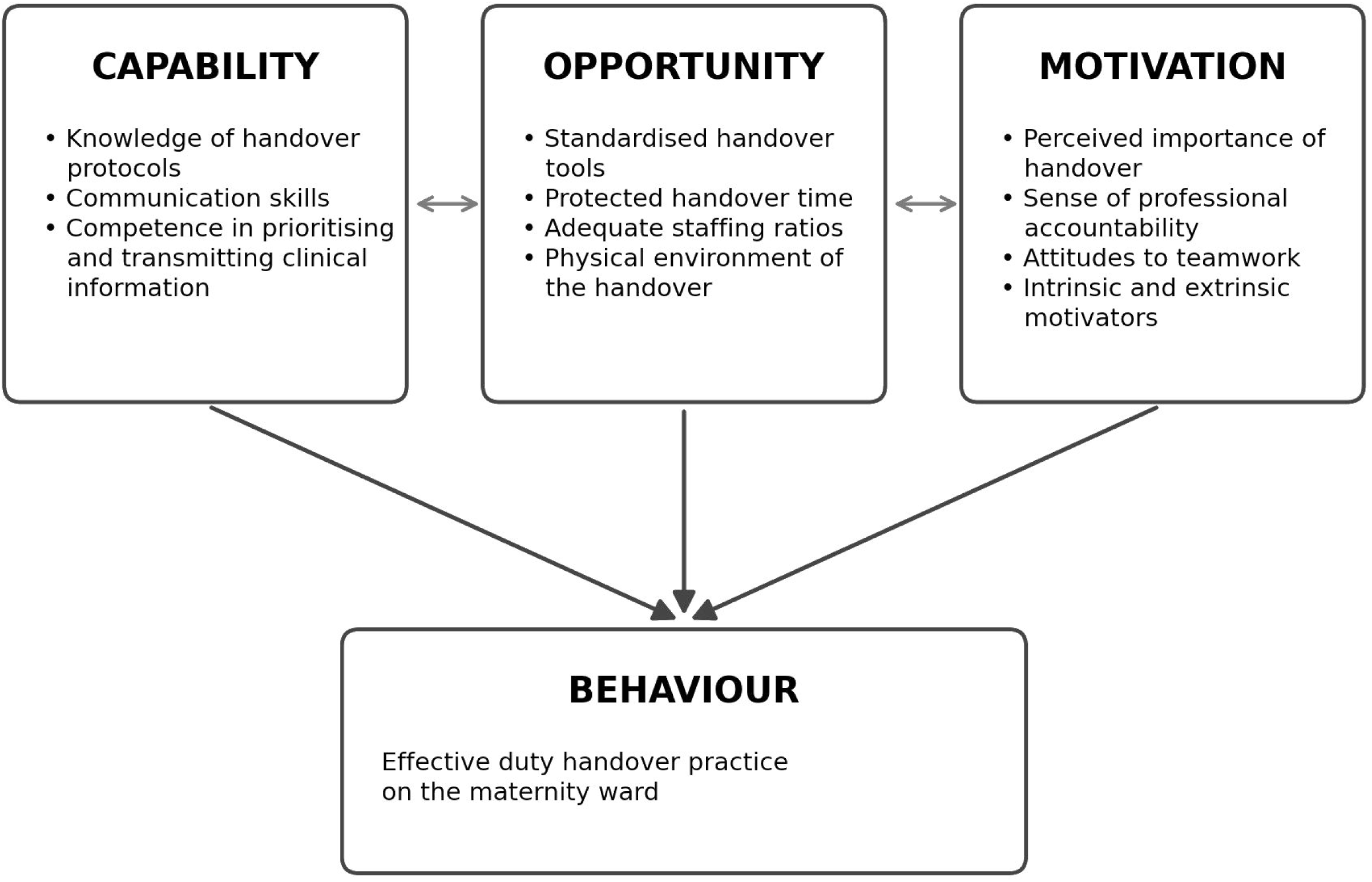
Conceptual framework: the COM-B model applied to duty handover practice on maternity wards. Capability, Opportunity and Motivation interact (double-headed arrows) and together influence Behaviour, here effective duty handover practice on the maternity ward. Adapted from Michie et al. [16].

### Researcher positionality

The corresponding author is an obstetrics and gynaecology registrar who had previously studied handover on the medical ward at HRRH in 2019, and who approached this study with a belief, formed through MPDSR reviews, that poor handover contributed to maternal and perinatal deaths.

This clinical and prior-research position gave him contextual familiarity with handover practice, but also created the potential for his assumptions to shape what he asked, how he probed and how he interpreted responses, particularly because he was a clinician interviewing midwives, nurses and clinical officers.

Reflexivity and bracketing were addressed in four stages. Before fieldwork, he wrote a dated statement of his prior knowledge and assumptions, covering his 2019 findings, his MPDSR-derived belief and his position as a registrar interviewing maternity staff. A senior author then held a bracketing interview with him on these assumptions, after which he revised the statement; both versions were kept. During data collection, he wrote a reflexive journal memo within 24 hours of each interview, recording what surprised him, what matched his expectations and where he noticed himself leading the participant. Field notes taken by research assistants were kept as a separate record. A senior author reviewed the journal after the fifth and fifteenth interviews and checked whether the interview guide or the principal investigator’s probing needed adjustment. During analysis, he reread his pre-fieldwork statement before opening any transcript. An independent coder (KSS) coded an agreed subset of transcripts without seeing the principal investigator’s codes, and the two coders met to compare codes and resolve differences. Any theme that closely matched the pre-study statement, or that mirrored a COM-B category, was tested against the transcripts for a counter-example, and the result was recorded. The journal, both versions of the pre-study statement and the coder comparison were retained by the research team. We acknowledge that bracketing can never be complete and that his position as an obstetrics registrar may have shaped what participants chose to say.

### Study setting

The study was conducted in public health facilities in Hoima District, in the Bunyoro sub-region of western Uganda: HRRH in Hoima municipality, one Health Centre IV and four Health Centre IIIs serving predominantly rural populations. Facilities were selected for their maternity case volumes, to represent different levels of the health system, and because they share the resource and staffing constraints that characterise public maternity care in Uganda. All facilities were within the public health system.

### Study population

The target population comprised maternity ward health workers employed in public health facilities in the Bunyoro sub-region and the wider western Uganda health system. The accessible population comprised maternity ward health workers and administrators employed in public health facilities in Hoima District at the time of the study. The study population comprised those who were available during the data collection period and gave voluntary written informed consent. Participants were eligible if they were trained maternity ward health workers or administrators who had served for at least 2 months in their current role in a public health facility in Hoima District. Those on leave, temporarily absent during data collection, or unavailable through severe illness or unforeseen circumstances were excluded.

### Sampling strategy

Purposive sampling with maximal variation was used to represent facility levels, professional cadres, experience and gender. Sample size was guided by saturation: Hennink and colleagues found that about nine interviews were sufficient to reach code saturation, whereas 16 to 24 were needed for meaning saturation [17]. We planned 30 participants (23 health workers and 7 administrators). Data collection stopped at 20 participants, comprising 15 health workers (in-depth interviews) and 5 administrators (key informant interviews), when interviews were no longer producing new concepts. The distribution of participants across facility levels is presented in the Results.

### Data collection

Participants were recruited between 10 March 2025 and 30 April 2025. Permission to access participants was obtained from the in-charges of the health centres and the Medical Superintendent of HRRH before enrolment. Prospective participants were approached individually, invited to participate voluntarily and briefed on the purpose of the study, the nature of their participation, the audio-recording procedures and the measures protecting their confidentiality; written informed consent was obtained before each interview. No participant declined or dropped out, and no one was interviewed more than once.

Data were collected by the principal investigator, with support from a co-author (JK) in data collection and management and from two trained research assistants with backgrounds in public health and the social sciences, who received two days of training in interviewing, ethics and study procedures. Interviews were conducted in English at the participating health facilities and lasted 10 to 30 minutes, depending on the respondent. Two research assistants were present during the interviews, in addition to the interviewer and the participant.

Separate semi-structured guides were used for in-depth interviews with health workers (12 questions) and key informant interviews with administrators (10 questions) (S1 File). Each guide covered the participant’s understanding of handover, available checklists and guidance, who hands over, when and how, effectiveness, barriers and suggestions for improvement. The guides contained open-ended questions aligned with the study objectives, with probes to deepen responses, and allowed flexibility to follow unanticipated lines of inquiry. They were pre-tested with four participants from facilities not included in the study to assess the clarity of questions, identify ambiguities and estimate interview duration, and were refined after daily debriefing sessions with the field team.

With permission, interviews were audio-recorded on a digital recorder and transcribed verbatim by research assistants, and the principal investigator checked each transcript against the recording. Research assistants also took field notes on non-verbal cues, context and reflections, which supplemented the audio data. Pseudonyms replaced participants’ names and recordings were stored securely. Interview guides were cross-checked daily for consistency and completeness, and a tracking system ensured that the number of guides distributed matched the number returned.

### Data analysis

Transcripts were analysed using a modified version of Colaizzi’s seven-step method [15]. The principal investigator read each transcript with the field notes to gain an overall sense of the data; extracted significant statements relevant to handover; formulated meanings from these statements; and clustered the formulated meanings into sub-themes and themes using a coding framework and coding matrix. An exhaustive description of the phenomenon was written from the themes, and its fundamental structure was summarised. Coding was done manually, without qualitative analysis software, by the principal investigator, with an independent coder (KSS) double-coding a subset of transcripts and an experienced qualitative researcher consulted on the coding framework; the research team discussed themes iteratively. Findings are presented as narrative themes supported by verbatim quotations identified by interview type (IDI, in-depth interview with a health worker; KII, key informant interview with an administrator).

The COM-B model [16] was applied after theme development as an interpretive lens, not as a deductive coding framework, so that themes remained grounded in participants’ accounts while the barriers could be organised by capability, opportunity and motivation. Themes that mirrored a COM-B category were tested against the transcripts for a counter-example.

### Rigour and trustworthiness

Credibility was supported by peer debriefing within the research team, repeated reading of the transcripts and the reflexivity procedures above. We did not return transcripts or findings to participants. Transferability was supported by sampling across facility levels, cadres and experience and by describing the setting in detail. Dependability was supported by documenting methodological decisions and instrument revisions throughout the study. Confirmability was supported by independent double-coding and by testing themes against the principal investigator’s pre-study assumptions. The study is reported in accordance with the Consolidated Criteria for Reporting Qualitative Research (COREQ) [18]; the completed checklist is provided as S2 File.

## Results

### Participant characteristics

Twenty participants were interviewed (15 health workers and 5 administrators), most of them women, reflecting the predominance of women in midwifery and nursing in Uganda’s public health system. The profile of participants by facility level is presented in **Table *1***. Participants were aged from 20 to above 40 years, with slightly more in the 20–40 age bracket, and their qualifications ranged from certificate to postgraduate level. Professional cadres included midwives, nurses, clinical officers, medical officers and administrators, and work experience ranged from under two years to more than five years, with a slight majority reporting more than five years of service.

**Table 1.** Profile of the 20 participants included in the study (n=20).

| Facility level | Participants (n) | Cadres represented | Range of experience | Gender | Age range |
| --- | --- | --- | --- | --- | --- |
| Hoima Regional Referral Hospital | 8 | Midwives, nurses, medical officers, administrators | <2 to >5 years | Male and female | 20–40 and above 40 years |
| Health Centre IV (1 facility) | 4 | Midwives, clinical officers, administrator | 2–5 and >5 years | Predominantly female | 20–40 and above 40 years |
| Health Centre III (4 facilities) | 8 | Midwives, nurses, clinical officers | <2 to >5 years | Mostly female, a few males | 20–40 and above 40 years |
| **Total** | **20** | **15 health workers; 5 administrators** |  |  |  |

### Overview of themes

Handover on the maternity wards varied considerably in structure, duration, standardisation and communication quality, and both formal and informal elements were present, with informal mechanisms predominating in several facilities. Two themes were identified, each with three sub-themes (**Table *2***), and are interpreted against the COM-B framework in the Discussion (**Fig *1***).

**Table 2.** Themes, sub-themes and codes describing duty handover practices and the challenges experienced by participants.

| Theme | Sub-theme | Codes |
| --- | --- | --- |
| 1. Duty handover practices on maternity wards | 1.1 Current state of duty handover | Existing handover protocol; handover duration and timing; information exchanged during handover |
|  | 1.2 Standardisation and consistency | Use of standardised handover tools; consistency in handover practices; adherence to established protocols |
|  | 1.3 Communication and collaboration | Effectiveness of communication during handover; collaboration between health care providers; involvement of patients and families |
| 2. Challenges experienced by health workers during the handover process | 2.1 Challenges in the handover process | Time constraints and workload; inadequate training or support; insufficient resources |
|  | 2.2 Communication breakdowns | Negative health worker attitudes; lack of clarity or competence in handover reports; difficulty accessing patient information |
|  | 2.3 Impact on patient care | Delays or errors in patient care; increased risk of adverse effects; impact on patient satisfaction and outcomes |

### Theme 1: Duty handover practices on maternity wards

#### Sub-theme 1.1: Current state of duty handover

Some form of handover protocol was acknowledged to exist in the facilities, but its implementation was largely informal and lacked a consistently applied, documented process (**Table *2***). The prevailing mechanism relied on telephone communication between outgoing and incoming staff rather than structured face-to-face reporting, a pattern that risks information loss and incomplete transfer of clinical responsibility.

> *“Their handover these days is more of a phone call — maybe midwife so-and-so says they will be leaving in like 30 minutes, and they find out whether the others will be available.” (KII, Administrator)*

The duration and timing of handover varied considerably between facilities and between shifts within the same facility. Several participants described handovers as brief and hurried, particularly when patient numbers were high.

> *“The duration of the handover is sometimes chap-chap. I will be direct to you — it even takes less than ten minutes.” (IDI, Health Worker)*

When handover was done properly, participants described the information exchanged as generally comprehensive, covering patients’ diagnoses, current conditions, ongoing treatment and notable clinical concerns. However, the quality of this exchange depended on individual initiative and professional commitment rather than on a protocol.

> *“I explain and describe all the patients that we have. I present them to the incoming doctor and explain how they are faring, the treatment they are receiving, and all the necessary information that would be beneficial for the patient and for the incoming doctor.” (IDI, Health Worker)*
>
> *“At maternity ward, we do give a report. We have both written and verbal reports. You give a report about the mothers who are in labour, how each mother is presenting, and which mother is a high-risk case.” (KII, Administrator)*

#### Sub-theme 1.2: Standardisation and consistency

Standardised handover tools were reported in some facilities but not in others. Where present, they gave the outgoing staff member a structured format for recording each patient’s status and clinical needs, which supported more consistent and complete information transfer. The variation in tool availability pointed to the absence of a unified, district-wide approach.

> *“The one who is leaving duty makes a written handover report in the handover tool, summarising the patients’ conditions and current state.” (KII, Administrator)*

Some participants reported a degree of consistency in handover practice within their facility and attributed it to teamwork, regular continuing medical education (CME) sessions and shared professional norms.

> *“There has been consistency, apparently, because there has been teamwork, conducting CMEs, and emphasising the most common obstetric practices.” (KII, Administrator)*
>
> *“At the end of the day, you realise that all of them are doing the same thing and working the same way, without bias and without attitude.” (IDI, Health Worker)*

Participants generally reported that providers adhered to the available protocols, completing critical tasks before the outgoing staff left and informing incoming staff of pending procedures and high-risk patients. Adherence was nevertheless described as uneven and dependent on individual commitment rather than on systematic oversight.

*“They are adhering to the protocols available here — to the best of my knowledge, they are not so bad.” (KII, Administrator)*

#### Sub-theme 1.3: Communication and collaboration

Communication during handover was described as more complete and reliable where structured protocols existed, and more prone to gaps where it relied on informal telephone contact. Most participants understood and practised the principle of finalising care for a patient before handing over, which reflected an awareness of the risks of transferring responsibility prematurely.

> *“In case there is any patient — like now, if I am monitoring a patient in labour — I am required first to finalise care for that patient before I hand over to another doctor.” (IDI, Health Worker)*

Participants also described instances of effective collaboration, in which outgoing and incoming staff exchanged information about patients, treatment plans and outstanding tasks. This was associated with smoother transitions and a shared sense of professional responsibility.

> *“When they come, they give a report. They tell each other how many patients are on the ward and what treatment each patient is receiving.” (IDI, Health Worker)*

Patients and families were generally not actively involved in handover. Where they knew about it, this arose from their own curiosity rather than from deliberate inclusion, which points to a gap in patient-centred care.

> *“Some patients would know — more so for the critically ill — and others may or may not know. If they do come to know, it will be out of curiosity.” (IDI, Health Worker)*

### Theme 2: Challenges experienced by health workers during the handover process

#### Sub-theme 2.1: Challenges in the handover process

Most participants named time constraints and workload as the greatest challenge. High patient numbers, poor time management and the late arrival of incoming staff compressed the time available, producing abbreviated verbal reports that lacked clinical depth (**Table *2***). This was most pronounced during periods of high obstetric activity, when comprehensive handover was most needed.

> *“The biggest challenge we have seen with midwives is some degree of poor time management. Some people take it lightly and come late, and they also find that when patient numbers are too many, you end up not giving a good handover report.” (KII, Administrator)*

Inadequate formal training in handover was described as a persistent barrier. Participants noted that many staff lacked the knowledge and skills for structured handover and voiced concern that the quality and professionalism of handover had declined over time. Recruitment that participants perceived to favour finances over technical competence was named as an aggravating factor.

> *“These days, handover of duty is not as smart as it used to be. People do their things to their will, and it is not systematic — maybe it is not professional.” (IDI, Health Worker)*
>
> *“There is still a knowledge gap, brought by people who are recruited into service based on finances other than technical ability.” (KII, Administrator)*

Participants also described a lack of supervisory oversight and accountability. The perceived decline in supervision was linked to less consistent and rigorous handover, because staff felt less accountable for the quality of their reports when they were not regularly monitored.

> *“There is less supervision at the moment. The health system these days is just like that — people care less about service delivery.” (IDI, Health Worker)*

#### Sub-theme 2.2: Communication breakdowns

Administrators and senior health workers identified negative attitudes among some staff as a significant impediment to handover. These attitudes took the form of disengagement from professional responsibilities and, in some cases, unwillingness to hand over before leaving a shift, which administrators attributed partly to informal recruitment and a resulting lack of professional commitment.

> *“We are managers, and we find it very hard when you cannot manage people like someone who paid their way into service. They don’t have the passion and the attitude towards duty handover and work in general.” (KII, Administrator)*
>
> *“Some people just don’t want to hand over. They think their duty is done, and they should go away. They just want to leave without handing over.” (KII, Administrator)*

Participants also raised concerns about the clarity and completeness of handover reports. Incomplete or insufficiently detailed reports left incoming staff without the information needed for safe and timely decisions, a problem compounded where written documentation was minimal and handover was verbal. Participants linked poor report quality to the knowledge gaps and limited training described above.

Incomplete patient records, disorganised documentation and limited availability of investigation results at handover were named as barriers to comprehensive reporting. Participants described situations in which missing vital signs, medications or laboratory results increased the risk of treatment errors.

> *“You find that you have not received the patient’s medical report very well — vitals like blood pressure and temperature, treatment given — you may find that a given treatment is not given in the right dose and time.” (IDI, Health Worker)*

#### Sub-theme 2.3: Impact on patient care

Participants linked inadequate handover directly to delays and errors in care. When critical information was not communicated, incoming staff could not prioritise care appropriately or respond promptly to deteriorating patients, which participants saw as especially worrying for the recognition of obstetric emergencies.

> *“Inadequate handover reports can lead to delays in care, as critical information may be missed or misunderstood.” (KII, Administrator)*
>
> *“When patient information is not communicated effectively, it can result in errors and delays in treatment, compromising patient safety.” (KII, Administrator)*

Participants also described poor communication at handover as contributing to adverse events, including medication errors, misdiagnosis and failure to recognise high-risk presentations, and emphasised that these failures increased patients’ vulnerability to preventable harm.

> *“Inadequate patient information and poor communication can increase the risk of adverse events and compromise patient safety.” (KII, Administrator)*
>
> *“When critical patient information is missing or not communicated effectively, it can lead to adverse outcomes and harm to patients.” (IDI, Health Worker)*

Finally, participants described consequences for patient satisfaction, including discontinuity of care, repeated unnecessary procedures and loss of trust arising from perceived carelessness.

> *“Inadequate care and poor communication can lead to decreased patient satisfaction and poor outcomes.” (IDI, Health Worker)*
>
> *“When patients don’t receive comprehensive care and support, it can affect their satisfaction and overall well-being.” (IDI, Health Worker)*

## Discussion

This study explored how health workers and administrators on maternity wards in Hoima District experience duty handover. Handover was inconsistent, relied heavily on informal telephone communication, and was constrained by limited time, training, resources and supervision, with participants linking these constraints to delays and errors in care (**Table *2***).

The predominance of informal, telephone-based handover is consistent with other African reports. In Kenyan newborn units, information exchange is verbal, formal reference documents are used rarely and ad hoc, and personal notes and notebooks are widespread [9,10]. In Ghanaian referral hospitals, the absence of formal handover procedures allowed shift teams to settle into default patterns of communication [5]. That participants in several facilities described the content of handover as depending on the individual rather than on a protocol (sub-themes 1.1 and 1.2) echoes the variation in handover content across staff and shifts reported in Kenya [9]. A general absence of formal handover policy and training, and separate midwifery and medical handovers, have also been described in Irish maternity services [11], so inconsistency is not confined to low-resource settings, although the absence of any enforced guideline appears to make it more pronounced here.

Where standardised tools were used, participants described more consistent and complete reports. Structured tools such as ISBAR are widely taught [7], a systematic review has examined structured handover interventions between nurses and patient safety outcomes [19], and an emergency-department study found that an ISBAR-based tool improved the completeness and accuracy of handover information [8]. Caution is nevertheless needed when transferring these tools to settings like Hoima. A pilot of a co-designed handover tool in Kenyan newborn units found that nurses valued it and perceived more complete information, but staff interest, supplies and high patient-to-staff ratios limited its use, and the authors suggested that such tools may work better as a communication guide than as an additional documentation burden [9]. This supports involving frontline staff in adapting any tool for Ugandan maternity wards. Participants’ emphasis on knowledge gaps also points to training: systematic reviews of educational interventions for handover and of teamwork and communication training support its inclusion in any improvement package [20,21].

Time pressure and workload, named by most participants, parallel findings elsewhere. Ghanaian nurses described taxing job demands that fed communication gaps [5], and in Kenyan newborn units, high patient-to-staff ratios and rushed, brief evening handovers with late-arriving staff limited how fully handover could be done [9]. Our participants’ accounts of late arrival and of reports shortened when patient numbers were high fit this pattern, and suggest that protected handover time and staffing are as important as the content of any tool.

Negative attitudes, including unwillingness to hand over, were described as a serious impediment. Poor work ethics and interpersonal conflict were among the attitudinal drivers of communication gaps in Ghana [5], and lack of interest and late arrival were reported as barriers in Kenya [9]. Difficulty accessing patient information resonates with a study in northern Tanzania that found incomplete medical records for surgical and obstetric patients [22], and with evidence that digital record systems in Sub-Saharan Africa are being developed and implemented but face substantial challenges [23]. Poor documentation was also a driver of communication gaps in Ghana [5]. Participants connected incomplete information to treatment errors; in this context, a survey of Ugandan health professionals suggested that medication errors are under-reported and that a non-punitive environment and patient involvement might encourage disclosure [24], which links safe handover to the wider safety culture of the facility.

Patients and families were largely absent from handover. Bedside approaches have been associated with greater patient participation and opportunities for patients to raise concerns [12], but participation in decision-making is only partly institutionalised in maternal and newborn health systems across LMICs [13]. Our findings suggest that inclusion of patients in handover in Hoima would require deliberate facility-level encouragement and adaptation to local circumstances, including privacy and the clinical state of women in labour and after birth, rather than simple transfer of high-income-country models.

### Factors that influenced handover guided by the COM-B model

Interpreting the findings through the COM-B model [16] (**Fig *1***; **Table *3***) showed that barriers in all three domains operated together, so that no single domain fully accounted for how handover was experienced.

**Table 3.** Barriers and enabling factors for duty handover reported by participants, organised by COM-B domain.

| COM-B domain | Barriers described by participants | Enabling factors described by participants |
| --- | --- | --- |
| Capability | Knowledge gaps and limited formal training in handover; incomplete or unclear reports; recruitment perceived to favour finances over technical competence | Continuing medical education sessions; shared professional norms; understanding of the need to finalise care before handing over |
| Opportunity | Time constraints, high patient numbers and late arrival of incoming staff; reliance on telephone handover; limited supervision; inconsistent availability of standardised tools; incomplete records and missing investigation results | Standardised written handover tools where available; combined written and verbal reports; teamwork among staff |
| Motivation | Negative attitudes; unwillingness to hand over before leaving a shift; weakened accountability where supervision was limited | Individual initiative and professional commitment; a shared sense of responsibility for patients |

COM-B, Capability, Opportunity, Motivation–Behaviour model.

Capability-related barriers included knowledge gaps and limited training in handover communication, which participants linked to incomplete or unclear handover reports and to the recruitment of staff perceived to lack technical competence.

Opportunity-related barriers included time constraints, workload, limited resources and supervision, inconsistent availability of standardised tools, and difficulty accessing patient information. These conditions shaped what even well-trained and well-motivated staff could achieve at the point of handover.

Motivation-related barriers included negative attitudes, unwillingness to hand over before leaving a shift, and weakened accountability where supervision was limited.

Because these barriers co-occur, tools or training addressing one domain alone may be insufficient. This is consistent with a multisite Australian study in which nursing handover improved when contextualised training was combined with changes to ward-level systemic barriers and was supported by ward leadership [25]. The barriers reported here also resemble those described by providers in public hospitals in India, where there was no formal handover system, little training and heavy patient loads [26]. The model therefore points towards a combined response of standardised tools adapted with frontline staff, targeted training, protected handover time and staffing, and stronger supervision and accountability.

### Strengths and limitations

Strengths include in-depth exploration of experience from both frontline workers and administrators across facility levels, a phenomenological design matched to the analytic method, and a structured, four-stage reflexivity and bracketing process that supports dependability and confirmability.

The study was confined to maternity wards in one district and its findings may not transfer to other departments, districts or private facilities, although we sought transferability through purposive sampling across facility levels and by describing the setting in detail. Participants may have described practice more favourably than it occurred (social desirability bias); we assured confidentiality and used probes, but cannot exclude this. The interviews relied on self-report and handover was not directly observed, so reported practices could not be verified. Interviews lasted 10 to 30 minutes and were held in English, which is the working language of the wards but may not be every participant’s first language, so some depth and nuance may have been lost. Transcripts and findings were not returned to participants for comment. Finally, the principal investigator’s prior clinical and research position on handover may have shaped which accounts were elicited and foregrounded despite the bracketing measures taken, and some participants may have been influenced by being interviewed by a registrar.

## Conclusions

Duty handover on the maternity wards studied in Hoima District was inconsistent in structure, duration and adherence to protocol, and relied heavily on informal telephone communication. Time constraints, inadequate training, limited resources and supervision, negative attitudes and poor access to patient information, which participants linked to delays and errors in care, operated together at the level of capability, opportunity and motivation. Patients and families were largely not involved in handover.

Practical priorities suggested by these findings are standardised handover tools adapted with frontline staff and supported by face-to-face, bedside handover complemented by written documentation; protected handover time and staffing; targeted training; stronger supervision, audit and accountability; and facility-and national-level guidance on handover for maternity care. Further research should evaluate the effect of standardised protocols and structured training on patient outcomes, develop and validate a locally adapted handover tool for Ugandan maternity wards, and explore handover from the perspective of women and families.

## Supporting information

Interview guides

COREQ: 32-item checklist

Human_Participants_Research_Checklist

somrec ethics approval letter

Administrative_Clearance_HRRH_Director_Stamped_20Mar2025

Administrative_Clearance_DHO_Health_Centres

## Author contributions

Conceptualisation: NT

Study design: NT, HK, MNK

Data collection and management: NT, JK

Data curation: NT, KSS

Formal analysis: NT, KSS, HK, MNK

Interpretation of findings: NT, HK, MNK

Writing – original draft preparation: NT, KSS

Writing – review and editing: NT, KSS, JK, HK, MNK

## Data availability statement

Interview transcripts describe named health facilities and professional roles within a small district workforce and cannot be shared publicly without risk of identifying participants, in line with the approved ethics protocol and the consent given. De-identified data are available on request, subject to approval by the Makerere University School of Medicine Research Ethics Committee (reference Mak-SOMREC-2024-1081), from: Makerere University College of Health Sciences, Department of Obstetrics and Gynaecology, P.O. Box 7072, Kampala, Uganda. The interview guides are provided as S1 File.

## Acknowledgments

We thank the health workers and administrators who took part in this study, the District Health Officer of Hoima District, the Hospital Director of Hoima Regional Referral Hospital and the in-charges of the participating health facilities for granting permission and facilitating access.

## Supporting information

**S1 File.** Interview guides. In-depth interview guide for maternity ward health workers and key informant interview guide for administrators.

**S2 File.** COREQ checklist. Consolidated Criteria for Reporting Qualitative Research (COREQ) 32-item checklist.

