## Supplementary material for "Health workers’ and administrators’ experiences of duty handover on maternity wards in public health facilities in Hoima District, Uganda: a qualitative phenomenological study": Interview guides

**S1 File. Interview guides**

### Appendix I: In-depth interview guide for maternity ward health workers

Number: ______________ Interviewer: ______________________________ Code: __________

Date of interview: ___________________

We are conducting a study assessing health workers' experience of duty handover on maternity wards in public health facilities in the Hoima district.

We will be asking you some questions; we request your responses about your experience, practices, challenges and recommendations as listed below.

1. What do you understand by duty handover on the maternity ward?
2. What do the Ministry of Health and the Government of Uganda recommend concerning duty handover in maternity wards?
3. Do you have any duty handover checklist on the maternity ward in this health facility?
4. How important is it to hand over duty on this ward?
5. Who does the handover of duty on the maternity ward in this health facility?
6. When is the handover of duty done on the maternity ward at this facility?
7. How is duty handover done on the maternity ward at this facility?
8. How effective is the handover of duty on the maternity ward at this health facility?
9. What barriers do you meet during the handover process in this health facility?
10. What do you think should be done to improve the handover of duty on the maternity ward in this health facility?
11. How does the health facility management support you to do handover duty well?
12. How does duty handover impact your day-to-day work on the maternity ward?

**Thank you for your time and active participation.**

### Appendix II: Key informant interview guide for administrators

13. What do you understand by duty handover on the maternity ward?
14. Do you have any handover checklist on the maternity ward in this health facility?
15. Who does the handover of duty on the maternity ward in this facility?
16. When is the handing over of duty done on the maternity ward in this facility?
17. How do maternity ward health workers perform duty handover in this health facility?
18. What do the Ministry of Health and the Government of Uganda recommend concerning duty handover on maternity wards?
19. Do you think health workers face some challenges during duty handover on the maternity ward in this health facility?
20. What are the benefits of duty handover on the maternity ward in this hospital?
21. How do you know if duty handover has been performed and vice versa on the maternity ward in this hospital?
22. What have you done to ensure that health workers on the maternity ward in this health facility observe effective handover of duty?

**Thank you for your time and active participation.**

*Both guides were approved by the Makerere University School of Medicine Research Ethics Committee (Mak-SOMREC-2024-1081) and are reproduced here as administered in the field, with minor formatting standardisation for this supporting file.*
