## Supplementary material for "Health workers’ and administrators’ experiences of duty handover on maternity wards in public health facilities in Hoima District, Uganda: a qualitative phenomenological study": COREQ: 32-item checklist

### S2 File. Consolidated criteria for reporting qualitative research (COREQ): 32-item checklist

Developed from: Tong A, Sainsbury P, Craig J. Consolidated criteria for reporting qualitative research (COREQ): a 32-item checklist for interviews and focus groups. Int J Qual Health Care. 2007;19(6):349–357. Line numbers refer to the line numbers in the submitted manuscript file.

| No. | Item | Guide question/description | Response | Reported on line(s) |
|---|---|---|---|---|
| **Domain 1: Research team and reflexivity** | | | | |
| ***Personal characteristics*** | | | | |
| 1 | **Interviewer/facilitator** | Which author/s conducted the interview or focus group? | Interviews were conducted by the principal investigator (NT, corresponding author), with support from a co-author (JK) in data collection and management and from two trained research assistants. | 179–182 |
| 2 | **Credentials** | What were the researcher's credentials? | The principal investigator is an obstetrics and gynaecology registrar (postgraduate trainee) in the Department of Obstetrics and Gynaecology, Makerere University. | 121–123 |
| 3 | **Occupation** | What was their occupation at the time of the study? | Obstetrics and gynaecology registrar; research assistants had backgrounds in public health and the social sciences. | 121–123; 179–181 |
| 4 | **Gender** | Was the researcher male or female? | The principal investigator is male (see Researcher positionality). | 121–127 |
| 5 | **Experience and training** | What experience or training did the researcher have? | Registrar with a prior study of handover on the medical ward at HRRH (2019). Research assistants received two days of training in interviewing, ethics and study procedures. | 121–123; 181–182 |
| ***Relationship with participants*** | | | | |
| 6 | **Relationship established** | Was a relationship established prior to study commencement? | No. Participants had no prior relationship with the interviewer or the research assistants. | — |
| 7 | **Participant knowledge of the interviewer** | What did the participants know about the researcher? | Participants were briefed on the study before consent; the participant information and consent form identified the principal investigator as a Master's student at Makerere University. | 174–177 |
| 8 | **Interviewer characteristics** | What characteristics were reported about the interviewer/facilitator? | Clinician-researcher with prior handover research and an MPDSR-derived belief that poor handover contributed to maternal and perinatal deaths; insider position and its possible influence are described. | 121–144 |
| **Domain 2: Study design** | | | | |
| ***Theoretical framework*** | | | | |
| 9 | **Methodological orientation and theory** | What methodological orientation was stated? | Qualitative phenomenological design; Colaizzi's descriptive phenomenological method for analysis; COM-B model used to interpret findings. | 102–106; 108–116; 201–206 |
| ***Participant selection*** | | | | |

| | | | | |
|---|---|---|---|---|
| 10 | **Sampling** | How were participants selected? | Purposive sampling with maximal variation (facility level, cadre, experience, gender). | 163–164 |
| 11 | **Method of approach** | How were participants approached? | Approached individually after permission from facility in-charges and the Medical Superintendent of HRRH, and invited to participate voluntarily. | 172–177 |
| 12 | **Sample size** | How many participants were in the study? | Twenty (15 health workers, 5 administrators); 30 were planned and recruitment stopped at 20 when no new concepts emerged. | 164–169; Table 1 |
| 13 | **Non-participation** | How many people refused to participate or dropped out? Reasons? | None. No participant refused to be interviewed and none dropped out; participants were purposively selected. | 177–178 |
| ***Setting*** | | | | |
| 14 | **Setting of data collection** | Where was the data collected? | Interviews were held at the participating public health facilities in Hoima District. | 146–151; 182–183 |
| 15 | **Presence of non-participants** | Was anyone else present besides the participants and researchers? | Yes. Two research assistants were present besides the participant and interviewer. | 183–184 |
| 16 | **Description of sample** | What are the important characteristics of the sample? | Facility level, cadre, experience, gender and age range are summarised in Table 1 and described in the Results. | 227–234; Table 1 |
| ***Data collection*** | | | | |
| 17 | **Interview guide** | Were questions, prompts, guides provided by the authors? Was it pilot tested? | Semi-structured guides (12-question IDI guide; 10-question KII guide) provided as S1 File; pre-tested with four participants from facilities not in the study. | 185–193 |
| 18 | **Repeat interviews** | Were repeat interviews carried out? | No. No participant was interviewed more than once. | 177–178 |
| 19 | **Audio/visual recording** | Did the research use audio or visual recording? | Yes. Interviews were audio-recorded with permission and transcribed verbatim. | 194–195 |
| 20 | **Field notes** | Were field notes made during and/or after the interview? | Yes. Research assistants took field notes on non-verbal cues, context and reflections; the principal investigator also kept a reflexive journal. | 132–134; 196–197 |
| 21 | **Duration** | What was the duration of the interviews? | Interviews lasted 10 to 30 minutes, depending on the respondent. | 182–183 |
| 22 | **Data saturation** | Was data saturation discussed? | Yes. Recruitment stopped at 20 participants when interviews no longer produced new concepts (informed by Hennink et al.). | 164–169 |
| 23 | **Transcripts returned** | Were transcripts returned to participants for comment and/or correction? | No. Transcripts were not returned to participants. | 218–219; 469–470 |
| **Domain 3: Analysis and findings** | | | | |
| ***Data analysis*** | | | | |
| 24 | **Number of data coders** | How many data coders coded the data? | The principal investigator coded all transcripts; an independent coder (KSS) double-coded an agreed subset; an experienced qualitative researcher was consulted. | 206–209 |
| 25 | **Description of the coding tree** | Did authors provide a description of the coding tree? | A coding framework and coding matrix were developed; themes, sub-themes and codes are shown in Table 2. | 201–206; Table 2 |

| | | | | |
|---|---|---|---|---|
| **26** | **Derivation of themes** | Were themes identified in advance or derived from the data? | Themes were derived from the data through Colaizzi's procedure; the COM-B model informed interpretation, and themes mirroring a COM-B category were tested for counter-examples. | 114–116; 139–141; 212–215 |
| **27** | **Software** | What software, if applicable, was used to manage the data? | No qualitative data analysis software was used; coding was done manually. | 206–209 |
| **28** | **Participant checking** | Did participants provide feedback on the findings? | No. Participants did not give feedback on the findings; findings were not returned to them (listed as a limitation). | 218–219; 469–470 |
| ***Reporting*** | | | | |
| **29** | **Quotations presented** | Were participant quotations presented to illustrate the themes? | Yes. Verbatim quotations are presented for each sub-theme and identified by interview type (IDI or KII). | 244–380 |
| **30** | **Data and findings consistent** | Was there consistency between the data presented and the findings? | Yes. Each sub-theme in Table 2 is illustrated by quotations in the Results and interpreted in the Discussion. | 244–380; 382–456 |
| **31** | **Clarity of major themes** | Were major themes clearly presented in the findings? | Yes. Two themes are presented with headings and an overview table. | 237–240 |
| **32** | **Clarity of minor themes** | Is there a description of diverse cases or discussion of minor themes? | Six sub-themes are described with illustrative quotations; limitations note the single-district scope and reliance on self-report. | 244–380; 462–473 |
