## Supplementary material for "Health workers’ and administrators’ experiences of duty handover on maternity wards in public health facilities in Hoima District, Uganda: a qualitative phenomenological study": somrec ethics approval letter

MAKERERE UNIVERSITY
P.O. Box 7072 Kampala, Uganda

COLLEGE OF HEALTH SCIENCES
SCHOOL OF MEDICINE
RESEARCH ETHICS COMMITTEE

**10/03/2025**

To: TUMWESIGE NICHOLAS

MAKERE UNIVERSITY KAMPALA
0782373880

**Type:** Initial Review

**Re: Mak-SOMREC-2024-1081: Health workers experience of duty handover on maternity ward at Hoima Regional Referral Hospital.**

I am pleased to inform you that at the **198** convened meeting on **10/12/2024**, the MAK School of Medicine REC (Mak-SOMREC) meeting voted to approve the above referenced application.
Approval of the research is for the period of **10/03/2025** to **10/03/2026.**

As Principal Investigator of the research, you are responsible for fulfilling the following requirements of approval:

1. All co-investigators must be kept informed of the status of the research.
2. Changes, amendments, and addenda to the protocol or the consent form must be submitted to the REC for re-review and approval **prior** to the activation of the changes.
3. Reports of unanticipated problems involving risks to participants or any new information which could change the risk benefit: ratio must be submitted to the REC.
4. Only approved consent forms are to be used in the enrollment of participants. All consent forms signed by participants and/or witnesses should be retained on file. The REC may conduct audits of all study records, and consent documentation may be part of such audits.
5. Continuing review application must be submitted to the REC **eight weeks** prior to the expiration date of **10/03/2026** in order to continue the study beyond the approved period. Failure to submit a continuing review application in a timely fashion may result in suspension or termination of the study.
6. The REC application number assigned to the research should be cited in any correspondence with the REC of record.
7. You are required to register the research protocol with the Uganda National Council for Science and Technology (UNCST) for final clearance to undertake the study in Uganda.

The following is the list of all documents approved in this application by MAK School of Medicine REC (Mak-SOMREC):
