## Supplementary material for "Health workers’ and administrators’ experiences of duty handover on maternity wards in public health facilities in Hoima District, Uganda: a qualitative phenomenological study": Administrative_Clearance_HRRH_Director_Stamped_20Mar2025

MAKERERE UNIVERSITY

P.O. Box 7072, Kampala, Uganda

**COLLEGE OF HEALTH SCIENCES**
**SCHOOL OF MEDICINE**
**DEPARTMENT OF OBSTETRICS AND GYNAECOLOGY**

Dr. Bwega Ibrahim

21st February 2025

To Whom It May Concern

HOSPITAL DIRECTOR
20 MAR 2025
P. O. BOX 5, HOIMA
HOIMA REGIONAL REFERRAL HOSPITAL

Dear Sir/Madam,

RE: **REQUEST TO CONDUCT RESEARCH IN YOUR FACILITY/DISTRICT**

This is to introduce to you Dr. Tumwesige Nicholas; Registration number 2021/HD07/3049U who is a postgraduate student in our department.

As part of his academic requirement, he will carry out research titled **"Health Worker's experience of duty hand over on maternity ward in Public Health facilities in Hoima district."**

Any assistance rendered to him will be highly appreciated.

Yours sincerely,

Dr Sekikubo Musa, MD; PhD
HEAD OF DEPARTMENT.
